# A mathematical model of COVID-19 transmission between frontliners and the general public

**DOI:** 10.1101/2020.03.27.20045195

**Authors:** Christian Alvin H. Buhat, Monica C. Torres, Yancee H. Olave, Maica Krizna A. Gavina, Edd Francis O. Felix, Gimelle B. Gamilla, Kyrell Vann B. Verano, Ariel L. Babierra, Jomar F. Rabajante

## Abstract

The number of COVID-19 cases is continuously increasing in different countries (as of March 2020) including the Philippines. It is estimated that the basic reproductive number of COVID-19 is around 1.5 to 4. The basic reproductive number characterizes the average number of persons that a primary case can directly infect in a population full of susceptible individuals. However, there can be superspreaders that can infect more than this estimated basic reproductive number. In this study, we formulate a conceptual mathematical model on the transmission dynamics of COVID-19 between the frontliners and the general public. We assume that the general public has a reproductive number between 1.5 to 4, and frontliners (e.g. healthcare workers, customer service and retail personnel, food service crews, and transport or delivery workers) have a higher reproduction number. Our simulations show that both the frontliners and the general public should be protected or resilient against the disease. Protecting only the frontliners will not result in flattening the epidemic curve. Protecting only the general public may flatten the epidemic curve but the infection risk faced by the frontliners is still high, which may eventually affect their work. Our simple model does not consider all factors involved in COVID-19 transmission in a community, but the insights from our model results remind us of the importance of community effort in controlling the transmission of the disease. All in all, the take-home message is that everyone in the community, whether a frontliner or not, should be protected or should implement preventive measures to avoid being infected.

## INTRODUCTION

The Coronavirus disease 2019 (COVID-19) poses a major global health threat. Several control measures are being done to minimize the spread of this contagious disease such as social distancing, case isolation, household quarantine, and school and university closure *[Ferguson, et. al 2020]*. Following China’s containment efforts, several countries adopted broad community quarantines or lockdowns as a means of controlling the spread of COVID-19 *[Anderson, 2020 and Cohen and Kupferschmidt 2020]*. However, amidst community quarantines and lockdowns, there are working classes who continue to provide essential services for healthcare, medicine, security, food, retail, and transport. This group of workers became collectively referred to as the “frontliners”. The nature of their work being in close proximity and in frequent interaction with the public puts them at a higher risk of getting infected *[Kiersz 2020 and Gamio 2020]*, and once infected, their continuous contact with the public can make them superspreaders. This led to the implementation of more stringent precautionary measures for frontliners especially to those working in healthcare.

In models of the spread of COVID-19, the key parameters are the basic reproduction number which refers to the average number of secondary cases generated from a contagious person, and a dispersion parameter that can provide further information about outbreak dynamics and potential for superspreading events *[Riou and Althaus 2020]*. It is estimated that the basic reproductive number of COVID-19 is around 1.5 to 4 *[Rabajante 2020]*. Parameters in models of disease spread are usually considered constant for the entire population with time-variation *[Liu, et.al. 2020]*. However, these parameters vary considering the heterogeneity of the population, location of virus transmission, and socio-economic and political factors *[Rabajante, 2020]*.

In this study, we formulate a mathematical model on the transmission dynamics of COVID-19 between the frontliners and the general public. We assume different basic reproductive numbers for the frontliners and the general public. We also consider a parameter for the susceptibility (which can be decreased through some level of protection) of an exposed individual with varying values for the frontliners and the general public. We examine the model simulations and perform sensitivity analysis to determine which parameters are influential to the model output.

## MATHEMATICAL MODEL

We consider an extended Susceptible-Exposed-Infected-Recovered (SEIR) Compartment Model to study the dynamics of the transmission of COVID-19 (Figure 1). The model has two mutually exclusive populations: the general public and the frontliners. Frontliners refer to working classes that provide continued services during disease outbreaks such as healthcare workers, customer service and retail personnel, food service crews, and transport or delivery workers.

**Figure 1.**
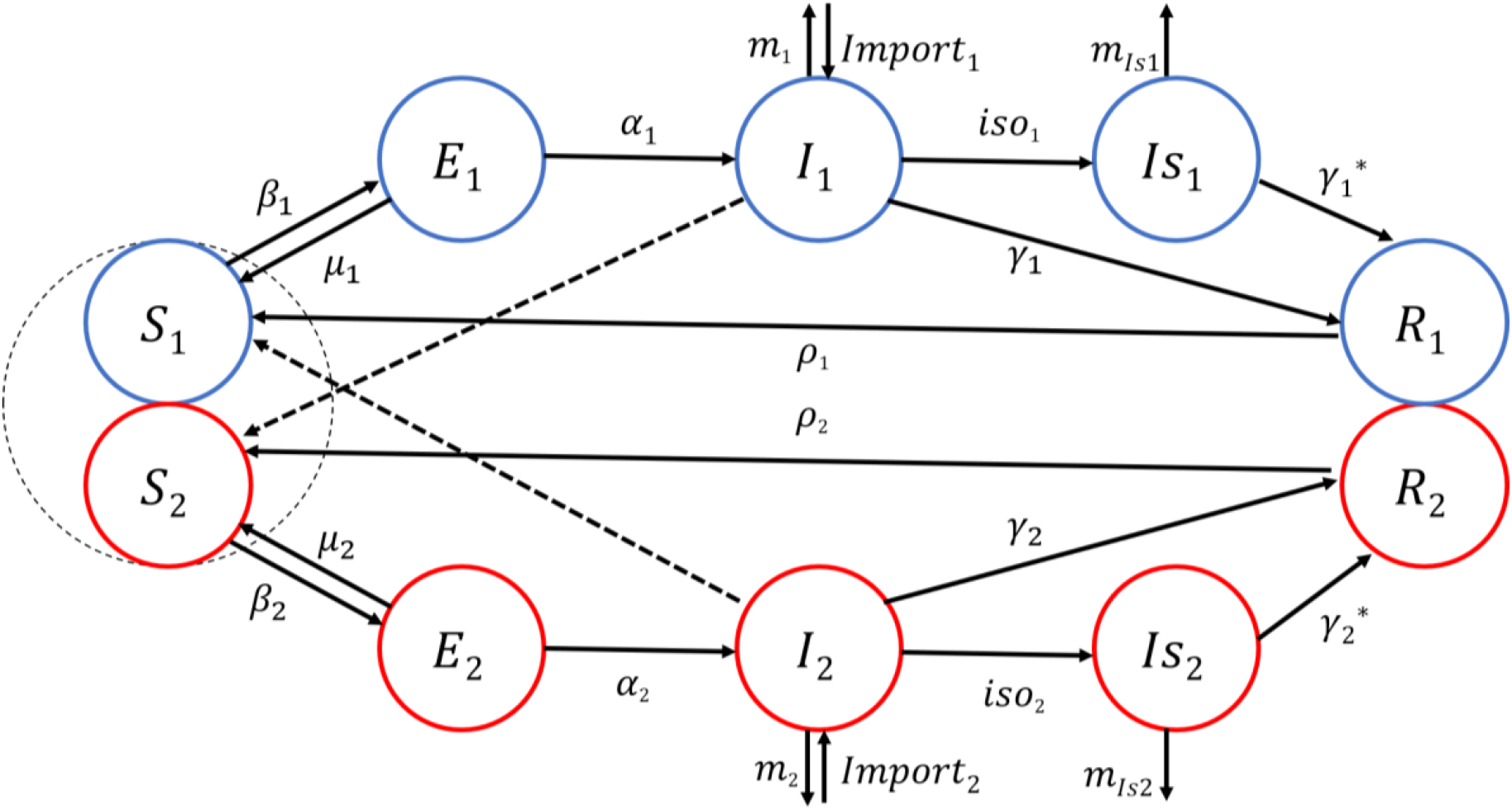
Extended Susceptible-Exposed-Infected-Recovered Model Framework of COVID-19 transmission between the general public and the frontliners. Two mutually exclusive populations with separate compartments for Susceptible, Exposed, Infected and Recovered are used to represent the dynamics of the transmission of the COVID-19 disease between these two populations.

In the model, the general public and the frontliners are compartmentalized to Susceptible, Exposed, Infected and Recovered. The numbers of susceptible public individuals and frontliners are *S*_1_ and *S*_2_, respectively. The numbers of individuals exposed to the disease are *E*_1_ for the general public and *E*_2_ for the frontliners. For the number of infected, *I*_1_ is for the general public and *I*_2_ is for the frontliners. From the infected, those that are in isolation are denoted by *Is*_1_ for the general public and *Is*_2_ for the frontliners. The numbers of recovered public individuals and frontliners are *R*_1_ and *R*_2_, respectively.

The general public and the frontliners are assigned different parameter values for basic reproduction number and susceptibility depending on the exposure to the disease. We refer to *β*_1_ and *β*_2_ as exposure rates for the general public and for the frontliners. The exposure rate is the number of new exposed individuals caused by an infectious individual per unit time. The rate at which an exposed public individual and an exposed frontliner become vulnerable or susceptible to the transmission of the virus is given by 1 − *μ*_1_ and 1 − *μ*_2_, respectively. Exposed individuals become infected with the disease at a rate of *α*_1_ for the general public and *α*_2_ for the frontliners. Moreover, an infected public individual is being isolated in a health facility at a rate of *iso*_1_ while an infected frontliner is being isolated at a rate of *iso*_2_. The rate of imported cases of infection is

*Import*_1_ for a public individual and *Import*_2_ for a frontliner. Without isolation, an infected public individual dies at a rate of *m*_1_ while an infected frontliner dies at a rate of *m*_2_. However, an infected public individual in isolation dies at a rate 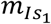 while for the frontliner, the rate is given by 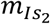. Without isolation, the rate of recovery is *γ*_1_ for an infected public individual and *γ*_2_ for an infected frontliner. On the other hand, in isolation, a public individual recovers at a rate of *γ*_1_^*\**^and a frontliner recovers at a rate of *γ*_2_^*\**^. Individuals who recovered from the disease become susceptible again at a rate of *ρ*_1_ for the general public and *ρ*_2_ for the frontliners.

The dynamics in the extended SEIR Compartment Model we used in the study is described by the following equations.

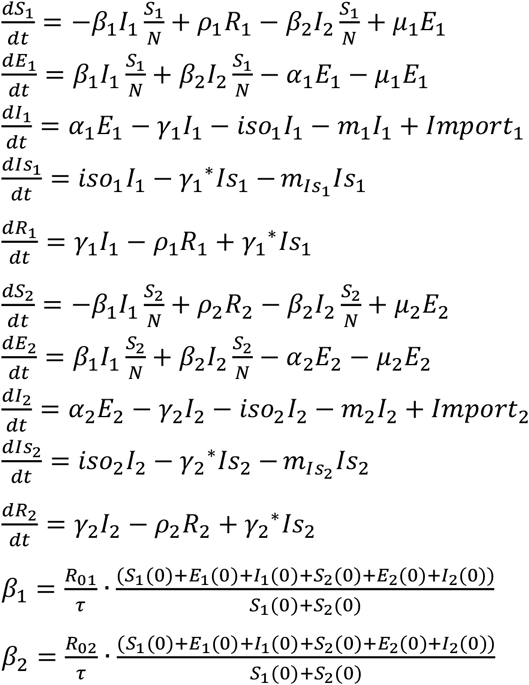

## SIMULATION RESULTS

For the simulation, we use parameter values indicated in Table 1. We run a simulation of the model for 200 days starting from the onset of the spread of the disease with initial values for population sizes indicated in Table 2. We vary parameter values for basic reproduction number and susceptibility (vulnerability) rate, and observe the number of infected individuals in isolation and not in isolation for both the frontliners and the general public. The following are our observations.

**Table 1:** Description of parameters and parameter values. The model parameters were estimated based on existing studies and observations on the current climate of the epidemic in the Philippines.

| Parameter | Description | Default Value | Reference |
| --- | --- | --- | --- |
| $R_{01}$ | Average number of secondary cases generated from a contagious public individual | 2.5 | [Anderson 2020] |
| $R_{02}$ | Average number of secondary cases generated from a contagious frontliner | 10 | Assumed |
| $\tau$ | Infectious period | 14 | [WHO 2020a] |
| $\beta_1$ | Exposure rate of a susceptible public individual | - | |
| $\beta_2$ | Exposure rate of a susceptible frontliner | - | |
| $1 - \mu_1$ | The rate at which an exposed public individual becomes susceptible or vulnerable to the transmission of the virus; $\mu_1$ is the average protection level of the public individuals | - | varied |
| $1 - \mu_2$ | The rate at which an exposed frontliner becomes susceptible or vulnerable to the transmission of the virus; $\mu_2$ is the average protection level of the frontliners | - | varied |
| $\alpha_1$ | Infection rate of an exposed public individual | 10/14 | [Rabajante 2020] |
| $\alpha_2$ | Infection rate of an exposed frontliner | 10/14 | [Rabajante 2020] |
| $iso_1$ | Probability of an infected public individual getting isolated to a health clinic | 0.01/14 | assumed |
| $iso_2$ | Probability of an infected frontliner getting isolated to a health clinic | 0.01/14 | assumed |
| $\gamma_1$ | Recovery rate of a non-isolated infected public individual | 0.96/14 | assumed |
| $\gamma_2$ | Recovery rate of a non-isolated infected frontliner | 0.96/14 | assumed |
| $\gamma^*_1$ | Recovery rate of an isolated infected public individual | 0.98/14 | assumed |
| $\gamma^*_2$ | Recovery rate of an isolated infected frontliner | 0.98/14 | assumed |
| $m_1$ | Death rate of an infected public individual | 0.03/14 | [Chen 2020] |
| $m_2$ | Death rate of an infected frontliner | 0.03/14 | [Chen 2020] |
| $m_{Is_1}$ | Death rate of an isolated infected public individual | 0.02/14 | assumed |
| $m_{Is_2}$ | Death rate of an isolated infected frontliner | 0.02/14 | assumed |
| $Import_1$ | Rate of imported cases of infected public individual | 0.1 | assumed |
| $Import_2$ | Rate of imported cases of infected frontliner | 0.1 | assumed |
| $\rho_1$ | Susceptibility rate of a recovered public individual | 0.1/30 | assumed |
| $\rho_2$ | Susceptibility rate of a recovered frontliner | 0.1/30 | assumed |

**Table 2.** Initial values for population of frontliners and the general public. To identify the effect on the dynamics of the initial value, we consider two sets of initial values for Susceptible, Exposed, Infected and Recovered both for the general public and the frontliners.

| Parameter | Description | Initial Value |  |
| --- | --- | --- | --- |
|  |  | Set 1 | Set 2 |
| $S_1$ | Number of susceptible from the general public | 10000 | 100000 |
| $S_2$ | Number of susceptible from the set of frontliners | 100 | 1000 |
| $E_1$ | Number of exposed from the general public | 0 | 0 |
| $E_2$ | Number of exposed from the frontliners | 10 | 100 |
| $I_1$ | Number of infected from the general public | 1 | 10 |
| $I_2$ | Number of infected from the set of frontliners | 0 | 0 |
| $Is_1$ | Number of infected in isolation from the general public | 0 | 0 |
| $Is_2$ | Number of infected in isolation from the set of frontliners | 0 | 0 |
| $R_1$ | Number of recovered from the general public | 0 | 0 |
| $R_2$ | Number of recovered from the set of frontliners | 0 | 0 |

The parameters *μ*_1_and *μ*_2_quantify the protection of the general public and the frontliners when they are exposed to the disease. Higher values for *μ* may mean that preventive measures (e.g. social distancing, use of protective gears and self-sanitizing) are effective against infection. We vary the values for *μ*_1_and *μ*_2_and observe how these affect the dynamics of the infection through the general public and the frontliners.

First, we increase the protection *μ*_1_of the general public, for a fixed value of *μ*_2_. We obtained the following insights: (i) as *μ*_1_ increases, there is a flattening in the peak of the number of infected for both populations (Figs. 2a-d), which means increasing the protection of the general public causes a significant decrease in the number of infected individuals; and (ii) as *μ*_1_ increases, the peak of the number of infected for both populations happens at a later time (Figs. 2), which implies the increase in the number of infected individuals is slowed down.

**Figure 2.**
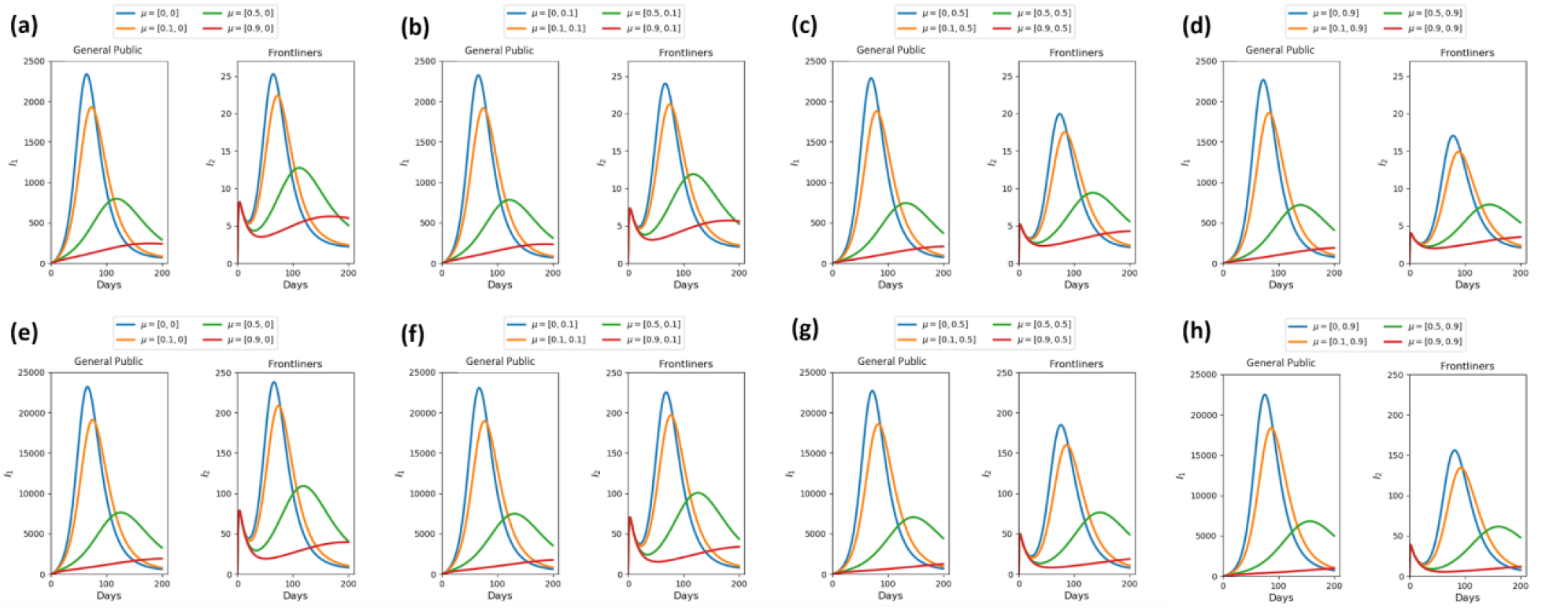
Predicted number of infected public individuals and frontliners when we increase the initial values of the compartments S, E, and I with fixed values of *μ*_2_and varying values of *μ*_1_. Parameter used: *R*_01_ = 2.5, *R*_02_ = 10,*τ* = 14, *iso*_1_ = *iso*_2_ = 0.01/14, *α*_1_ = *α*_2_ = 10/14, *γ*_1_ = *γ*_2_ = 0.96/14,*γ*_1_ *\** = *γ*_2_ *\** = 0.98/14, *m*_1_ = *m*_2_ = 0.03/14, *m*_*Is*1_ = *m*_*Is*2_ = 0.02/14, *Import*_1_ = *Import*_2_ = 0.1. (**a-d**) Initial population *S*_1_ = 10000, *E*_1_ = 0, *I*_1_ = 1, *Is*_1_ = 0, *R*_1_ = 0, *S*_2_ = 100, *E*_2_ = 10, *I*_2_ = 0, *Is*_2_ = 0, *R*_2_ = 0. (**e-h**) Initial population *S*_1_ = 100000, *E*_1_ = 0, *I*_1_ = 10, *Is*_1_ = 0, *R*_1_ = 0, *S*_2_ = 1000, *E*_2_ = 100, *I*_2_ = 0, *Is*_2_ = 0, *R*_2_ = 0. (**a-h**) *μ*_1_ = 0, 0.1,0.5, 0.9. (**a**,**e**) *μ*_2_ = 0. (**b**,**f**) *μ*_2_ = 0.1. (**c**,**g**) *μ*_2_ = 0.5. (**d**,**h**) *μ*_2_ = 0.9.

Second, we significantly change the initial values of the compartments S,E, and I but with varying values of both *μ*_1_and *μ*_2_(Fig. 3). We observed that peaks are reached after almost the same number of days regardless of the initial population size.

**Figure 3.**
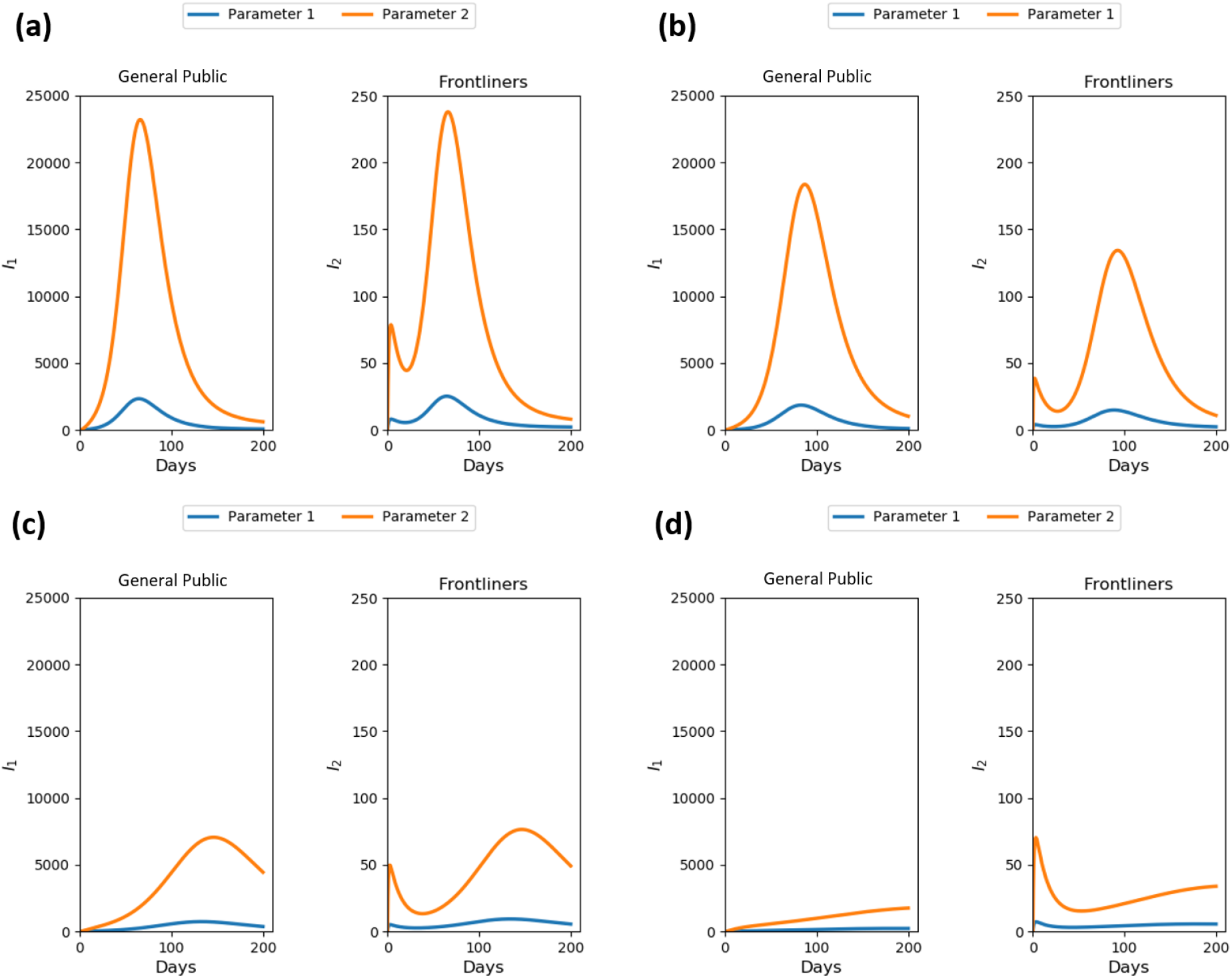
Predicted number of infected public individuals and frontliners when we increase the initial values of the compartments S, E, and I with fixed values of *μ*_1_and *μ*_2_. Parameter used: *R*_01_ = 2.5, *R*_02_ = 10,*τ* = 14, *iso*_1_ = *iso*_2_ = 0.01/14, *α*_1_ = *α*_2_ = 10/14, *γ*_1_ = *γ*_2_ = 0.96/ 14,*γ*_1_ *\** = *γ*_2_ *\** = 0.98/14, *m*_1_ = *m*_2_ = 0.03/14, *m*_*Is*1_ = *m*_*Is*2_ = 0.02/14. (**Parameter 1**) Initial population *S*_1_ = 10000, *E*_1_ = 0, *I*_1_ = 1, *Is*_1_ = 0, *R*_1_ = 0, *S*_2_ = 100, *E*_2_ = 10, *I*_2_ = 0, *Is*_2_ = 0, *R*_2_ = 0. (**Parameter 2**) Initial population *S*_1_ = 100000, *E*_1_ = 0, *I*_1_ = 10, *Is*_1_ = 0, *R*_1_ = 0, *S*_2_ = 1000, *E*_2_ = 100, *I*_2_ = 0, *Is*_2_ = 0, *R*_2_ = 0. (**a**) *μ*_1_ = 0, *μ*_2_ = 0. (**b**) *μ*_1_ = 0.1, *μ*_2_ = 0.9. (**c**) *μ*_1_ = 0.5, *μ*_2_ = 0.5. (**d**) *μ*_1_ = 0.9, *μ*_2_ = 0.1.

Third, we increase the protection *μ*_2_of the frontliners, for a fixed value of *μ*_1_. The number of infected frontliners decreased but there is no significant effect on the number of infected public individuals (Fig. 4). Moreover, when the initial population of frontliners increased ten times, the peak on the number of infected public individuals significantly increased by 43.48% of the total number of susceptible public individuals. When we increase the protection of the frontliners, we observe a decrease in the number of infected public individuals. We also found that decreasing the average secondary infections produced by an infectious public individual (*R*_01_) will considerably reduce the infected population of both the general public and the frontliners (Fig.5a). Figure 5b shows that the most effective way to minimize the number of infected individuals is a combination of reduced reproductive number (*R*_01_) and an improved protection for the general public. Moreover, when the average number of secondary cases generated from a contagious public individual and a contagious frontliner is 1.5 and 2.5, respectively (i.e., *R*_01_ = 1.5 and *R*_02_= 2.5), there is only a small chance that the general public will infect the frontliners (Fig. 5), thus protecting the frontliners. This agrees with the measures currently being implemented that the general public should practice social distancing and be protected.

**Figure 4.**
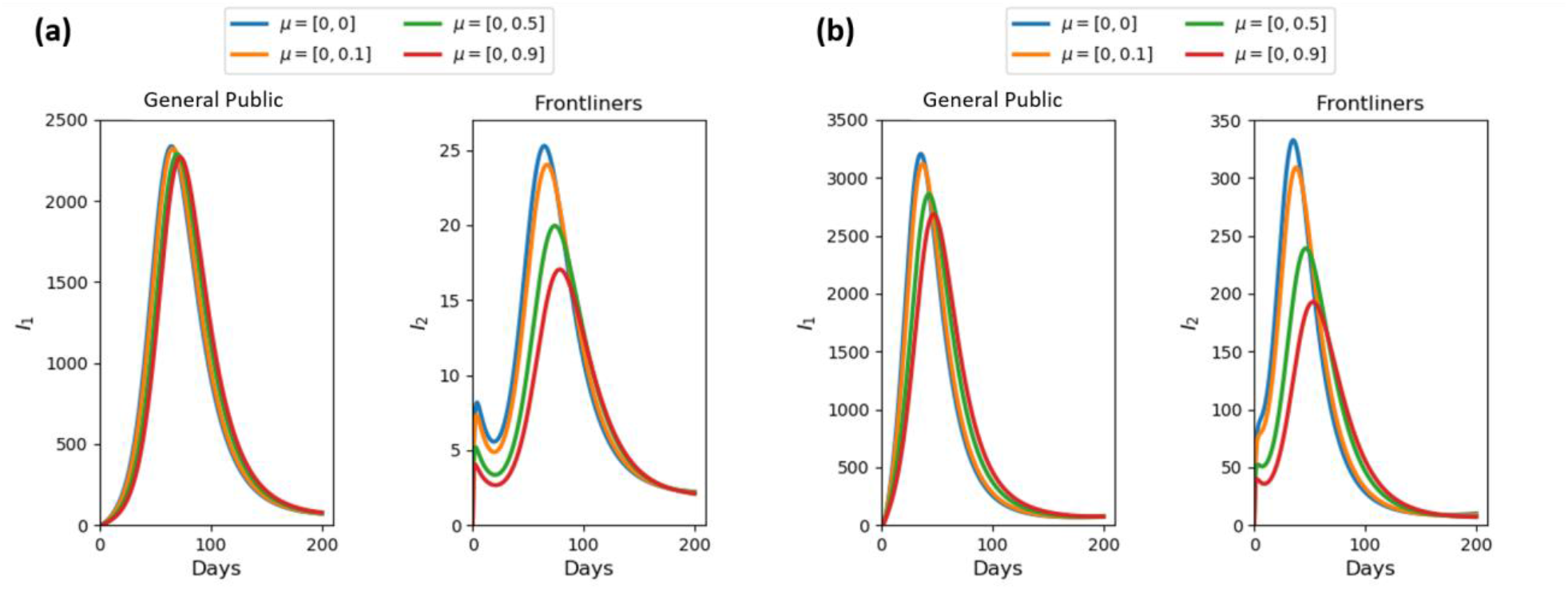
Predicted number of infected public individuals and frontliners when we increase the initial values of the compartments S, E, and I of the frontliners ten times with fixed values of *μ*_1_and varying values of *μ*_2_. Parameter used: *S*_1_ = 10000, *E*_1_ = 0, *I*_1_ = 1, *Is*_1_ = 0, *R*_1_ = 0, *R*_01_ = 2.5, *R*_02_ = 10,*τ* = 14, *iso*_1_ = *iso*_2_ = 0.01/14, *α*_1_ = *α*_2_ = 10/14, *γ*_1_ = *γ*_2_ = 0.96/14,*γ*_1_ *\** = *γ*_2_ *\** = 0.98/14, *m*_1_ = *m*_2_ = 0.03/14, *m*_*Is*1_ = *m*_*Is*2_ = 0.02/14. (**a**) Initial population *S*_2_ = 100, *E*_2_ = 10, *I*_2_ = 0, *Is*_2_ = 0, *R*_2_ = 0. (**b**) Initial population *S*_2_ = 1000, *E*_2_ = 100, *I*_2_ = 0, *Is*_2_ = 0, *R*_2_ = 0. (**a-b)** *μ*_1_ = 0, *μ*_2_ = 0,0,0.1,0.5,0.9.

**Figure 5.**
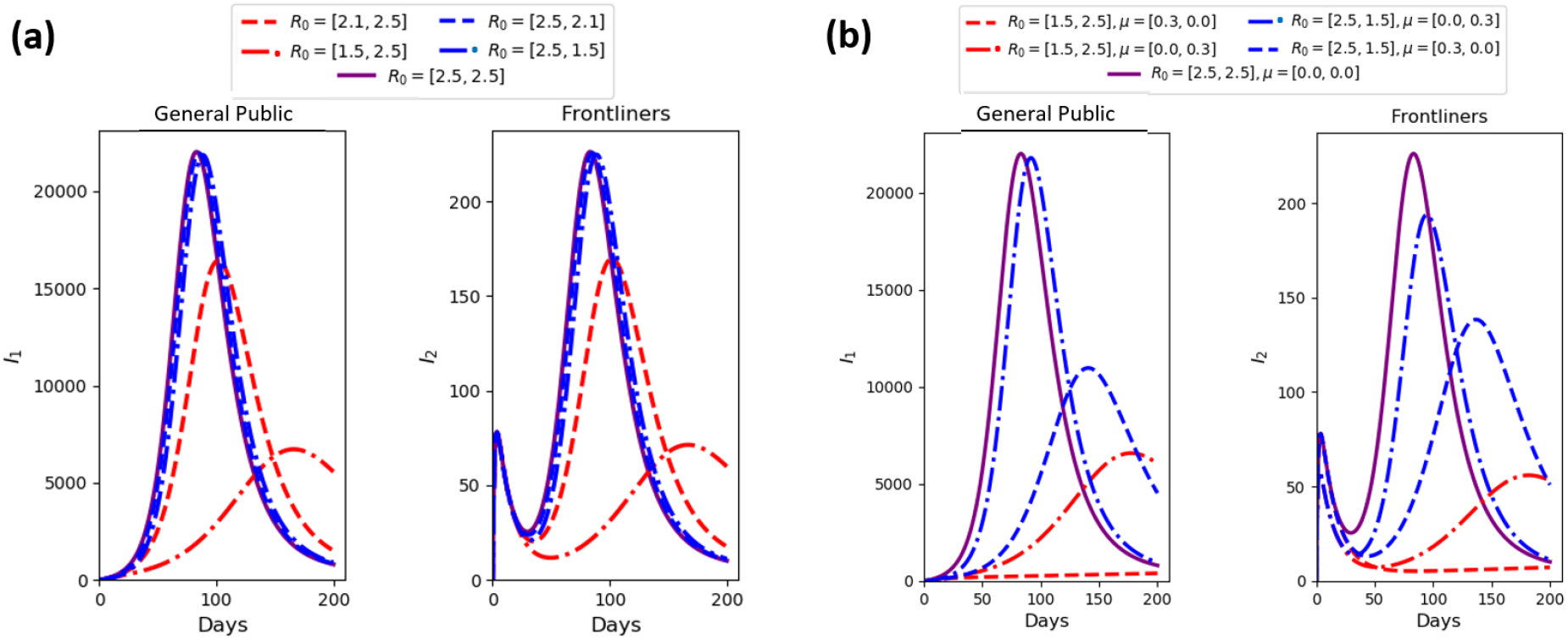
Predicted number of infected public individuals and frontliners when the level of protection varies. (**a**) Number of infected when we vary *R*_0_ with fixed values of *μ*_1_and *μ*_2_. (**b**) Number of infected when we vary *R*_0_, *μ*_1_and *μ*_2_. Parameter used: Initial population *S*_1_ = 100000, *E*_1_ = 0, *I*_1_ = 10, *Is*_1_ = 0, *R*_1_ = 0, *S*_2_ = 1000, *E*_2_ = 100, *I*_2_ = 0, *Is*_2_ = 0, *R*_2_ = 0,*R*_01_ = 2.5, *R*_02_ = 10,*τ* = 14, *iso*_1_ = *iso*_2_ = 0.01/14, *α*_1_ = *α*_2_ = 10/14, *γ*_1_ = *γ*_2_ = 0.96/14,*γ*_1_ *\** = *γ*_2_ *\** = 0.98/14, *m*_1_ = *m*_2_ = 0.03/14, *m*_*Is*1_ = *m*_*Is*2_ = 0.02/14. (**a**) *μ*_1_ = 0, *μ*_2_ = 0.

## SENSITIVITY ANALYSIS

Sensitivity analysis is a method used to identify the effect of each parameter in the model outcome. Its aim is to identify the parameters that most influence the model output and quantify how uncertainty in the input affects model outputs [*Marino et al*., *2008*]. In this study, we are interested in the number of infected individuals of both the general public *I*_1_ and the frontliners *I*_2_. We employed a method called partial rank correlation coefficient (PRCC) analysis, which is a global sensitivity analysis technique that is proven to be the most reliable and efficient sampling-based method. To implement PRCC analysis, Latin Hypercube Sampling (LHS) is used in obtaining input parameter values. This is a stratified sampling without replacement technique proposed by Mckay et al. [*Marino et al*., *2008*]. Here, the uniform distribution is assigned to every parameter and simulation is done 10,000 times. The maximum and minimum values of the parameters are set as ±90% of the default values listed in Table 1 with values of *μ*_1_ and *μ*_2_ set to 0.1.

PRCC values which range from -1 to 1 are computed in different time points, specifically in the years t=40k, k=0,1…,5 using the MATLAB function partialcorr. In Figure 6, each bar corresponds to a PRCC value at an instance. The value of 1 takes a perfect positive linear relationship while -1 means a perfect negative linear relationship, and a large absolute PRCC value would mean a large correlation of the parameter with the model outcome, that is, a minute change to a sensitive parameter would affect the dynamics of the model output. Parameters *R*_01_,*τ, α*_1_, *γ*_1_, and *R*_02_ are found to have high PRCC values (>0.5 or <-0.5). Among these, *R*_01_, *α*_1_, and *R*_02_ have positive PRCC values which mean that an increase in the values of these parameters will result in an increase in the infected population size. In contrast, *τ* and *γ*_1_ have negative PRCC values which indicate that increasing their values will consequently decrease the infected population.

**Figure 6.**
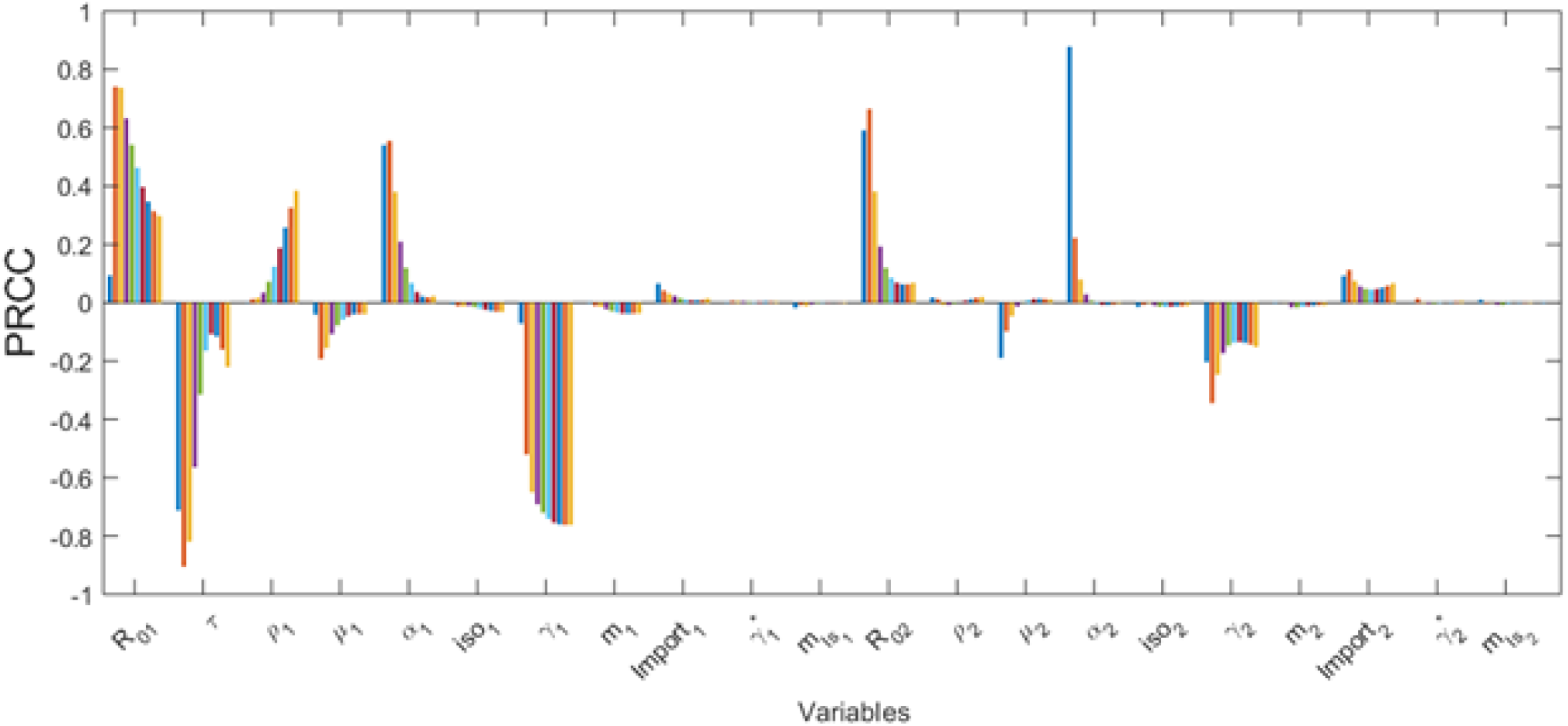
PRCC values depicting the sensitivities of the model output infected population with respect to model parameters.

## DISCUSSION

We investigated the transmission dynamics of COVID-19 between frontliners and the general public using an extended Susceptible-Exposed-Infected-Recovered (SEIR) Compartment Model. The model has two mutually exclusive populations: the general public and the frontliners. Since frontliners have frequent interaction with the population, we assume a basic reproduction number higher than that of the general public. Sensitivity of the model parameters is determined to evaluate those with significant impact on the model output, in this case, the infected population of both the general public and frontliners. It was observed that the infected population is sensitive to the changes in the basic reproduction numbers of the general public *R*_01_ and of the frontliners *R*_02_, the infection period *τ*, the infection rate of an exposed public individual *α*_1_, and the recovery rate of a non-isolated infected public individual *γ*_1_.

The model cannot be immediately utilized to make predictions on the spread of COVID-19 but it provides us insights on the transmission of a disease between two populations with different characteristics in terms of factors affecting the spread of a disease, such as the basic reproduction number and susceptibility rate. This can help in developing sound decisions and effective strategies in mitigating the spread of a disease. Simulations of the model show that both the frontliners and the general public should be protected or resilient against a spreading disease. Prioritizing only the protection of the frontliners cannot flatten the epidemic curve. On the other hand, protecting only the general public from the disease will significantly flatten the epidemic curve but the infection risk faced by the frontliners is still high, which can eventually affect their capability to provide services during an epidemic. In addition, if the control measures for the public are less strict, we can expect that the number of secondary cases to be higher on the average.

## Data Availability

Code available upon request to authors

